# Efficacy of Spirulina for Allergic Rhinitis

**DOI:** 10.1101/2021.10.21.21265199

**Authors:** Iury Gomes Batista, Osmar Clayton Person, Priscila Bogar, Fernando Veiga Angélico Júnior

## Abstract

**Introduction:** Allergic rhinitis is a condition of high prevalence in the population and widely studied, with several treatments being consecrated for its control. Spirulina is a dietary supplement that modulates immune function, and has been shown to modulate the inflammatory response of allergic rhinitis.

**Purpose:** To evaluate spirulina in the treatment and control of allergic rhinitis.

**Material and Methods:** This is a systematic review of randomized clinical trials. Searches were performed for randomized clinical trials relating spirulina to allergic rhinitis in five electronic databases: Cochrane - Central Register of Controlled Trials - CENTRAL (2021), PUBMED (1966-2021), EMBASE (1974-2021), LILACS (1982-2021) AND SCOPUS (2021). Two investigators independently extracted data and assessed trial quality.

**Results:** Two clinical trials involving a total of 215 patients were included. Both studies assessed the efficacy of spirulina in improving allergic rhinitis as the primary outcome. The first study described a significant reduction in runny nose, nasal congestion and itching over time of medication use (p<0.001) and in the second study the prevalence of rhinorrhea (P = 0.021), nasal congestion or obstruction (P = 0.039) and decreased smell (P = 0.030) were significantly less in the experimental group than in the control group.

**Conclusions:** The included studies were in favor of the use of spirrulina. However, the level of evidence is very low and limited. We should have caution due to the small number of clinical trials and participants in these studies. It is recommended to carry out new RCTs following the CONSORT standardization.

## INTRODUCTION

Allergic rhinitis (AR) is an atopic disease characterized by symptoms of nasal congestion, rhinorrhea, sneezing, post-nasal drip and itchy nose. It affects one in six individuals and is associated with significant morbidity, lost productivity and health care costs. (1)

The prevalence of allergic rhinitis is approximately 15%; however, the prevalence is estimated to be up to 30% based on patients with nasal symptoms. AR is known to peak in the second to fourth decades of life and then gradually decline. The incidence of AR in the pediatric population is also quite high, making it one of the most common chronic pediatric diseases. (two)

The allergic response is divided into two phases: early and late. In the early stage, allergic rhinitis is mediated by immunoglobulin IgE against inhaled allergens that cause inflammation induced by type 2 helper cells (Th2). (3) The initial response occurs within 5 to 15 minutes after exposure to an antigen, resulting in degranulation of the host’s mast cells. This releases a variety of preformed and newly synthesized mediators, including histamine, which is one of the primary mediators of allergic rhinitis. Histamine induces sneezing through the trigeminal nerve and also plays a role in rhinorrhea by stimulating the mucous glands. Other immunological mediators, such as leukotrienes and prostaglandins, are also implicated, as they act on blood vessels to cause nasal congestion. Four to six hours after the initial response, there is an influx of cytokines, such as interleukins (IL)-4 and IL-13 from mast cells, signifying the development of a late-phase response. These cytokines, in turn, facilitate the infiltration of eosinophils, T lymphocytes and basophils in the nasal mucosa and produce nasal edema with resulting congestion (4).

Risk factors for developing AR include family history of atopy, male gender, presence of allergen-specific IgE, serum IgE greater than 100 IU/mL before 6 years of age, and higher socioeconomic status. (5)

In the treatment of AR, there are several pharmacological options used to control and treat symptoms, the main options being: antihistamines, intranasal corticosteroids, leukotriene receptor antagonists) and immunotherapy. Even with so many options, AR still remains inadequately controlled with current means, so new classes of drugs are emerging every day for its treatment. The need for continuous drug therapy makes individuals anxious about the long-term effects of drugs. Therefore, an alternative strategy is needed. (6)

Effects of spirulina and tinospora cordifolis were investigated on allergic rhinitis. (6) Spirulina (Aphanizomenon sp., Spirulina sp.) is a type of cyanobacterium (blue-green freshwater algae) that contains various nutrients, such as proteins, B vitamins, vitamin E, chlorophyll, beta-carotene and iron. Its use dates back to the 16th century, when the Aztec peoples consumed it as part of their diet. (7)

A number of studies have suggested that spirulina is an effective modulator of the immune system. It is well documented in the literature that spirulina exhibited anti-inflammatory properties, in particular, by inhibiting histamine release from mast cell-mediated allergics. (8) The active ingredient found in spirulina responsible for its anti-inflammatory activities is C-phycocyanin, a pigment commonly found in blue-green algae. It is known that C-phycocyanin can selectively inhibit the activity of cyclooxygenase-2, an enzyme responsible for prostaglandin biosynthesis. Because of its chemical structure, C-phycocyanin also exhibits antioxidant and free radical scavenging properties, which may contribute, at least in part, to its anti-inflammatory activities. More recently, we showed that the in vitro culture of spirulina extract was effective in modulating the secretion of cytokines [interleukin (IL)-1, IL-4 and interferon (IFN). (9, 10.11)

## PURPOSE

The study aimed to evaluate the effectiveness of the use of spirulina in the treatment of allergic rhinitis, especially with regard to:

- the subjective clinical improvement of the symptom;
- the improvement in the quality of life;

## MATERIAL AND METHODS

This is a systematic review of randomized clinical trials, following the methodology recommended by the Cochrane Collaboration (12), carried out in the Discipline of Otorhinolaryngology, Faculty of Medicine of ABC (FMABC).

Only randomized controlled trials (RCTs) were included in the study, whose participants were adults and children of both sexes with allergic rhinitis, regardless of symptom severity and duration.

### Types of Interventions

Spirulina-treated group, with a dosage of 2g/day for a period of 24 weeks, compared to the placebo-treated group.

Group treated with spirulina, with a dosage of 2g/day for a period of 12 weeks, compared to the group treated with cetirizine 10mg/day.

### Types of Outcomes

- Primary
  - Subjective improvement of allergic rhinitis symptoms

- Secondary
  - Improved quality of life

### Search Methods for Identification of Studies

The search strategy developed was recommended by chapter six of the Cochrane Collaboration Handbook, a high-sensitivity search strategy. The keywords “allergic rhinitis” and “spirulina” were used, also using the Cochrane filter for study identification (ECR).

The official vocabulary identified was extracted from DECS – Descriptor in Health Sciences – http://decs.bvs.br/ and MeSH – Medical Subject Headings – http://www.ncbi.nlm.nih.gov/mesh. The following descriptors and terms were used: “Rhinitis, Allergic”[Mesh] AND Spirulina [Mesh}. No filter was used to identify specific study designs.

Five electronic databases were searched: Cochrane Library - CENTRAL (2021), MEDLINE/PUBMED (1966-2021), EMBASE (1974-2021), LILACS (1982-2021) and SCOPUS (2021). The date of the last survey was June 3, 2021. There were no restrictions on the language or geographic origin of the publications.

The search strategy for researching and identifying studies in electronic databases is shown in table 1. Only randomized clinical trials (RCT) were selected in the study.

**Table 1:**
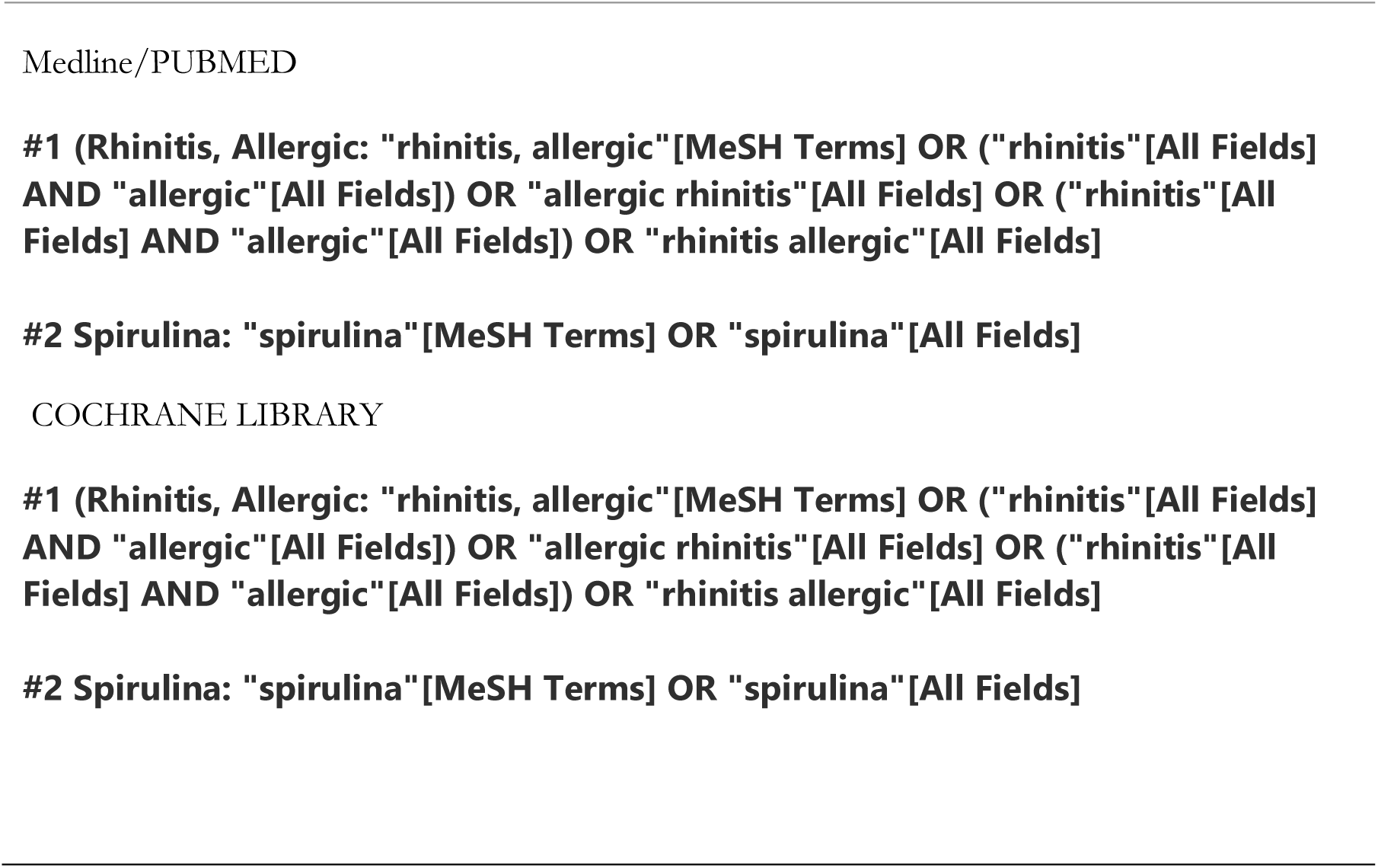
Search strategy for Medline and Cochrane databases.

**Table 1.**
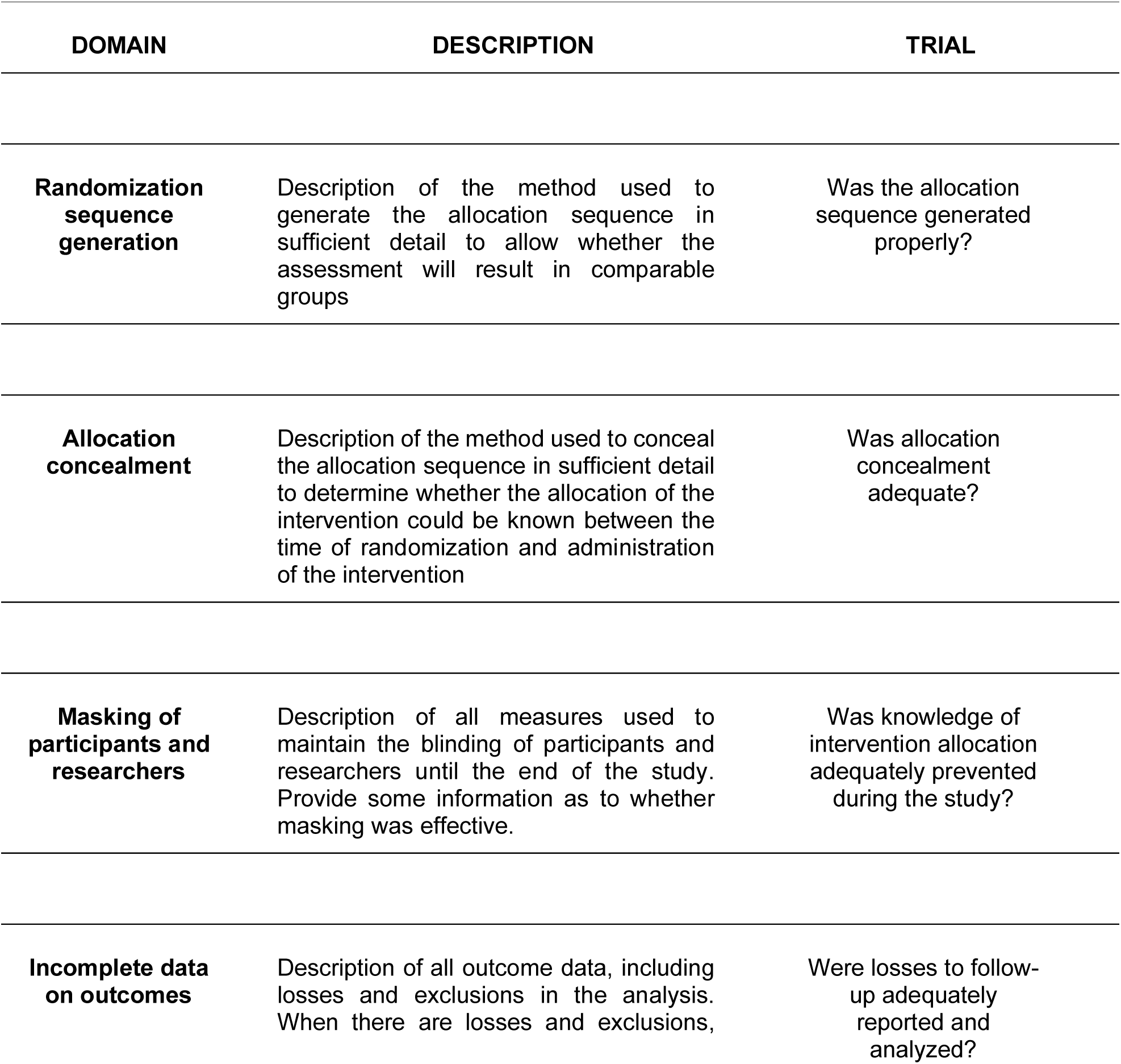
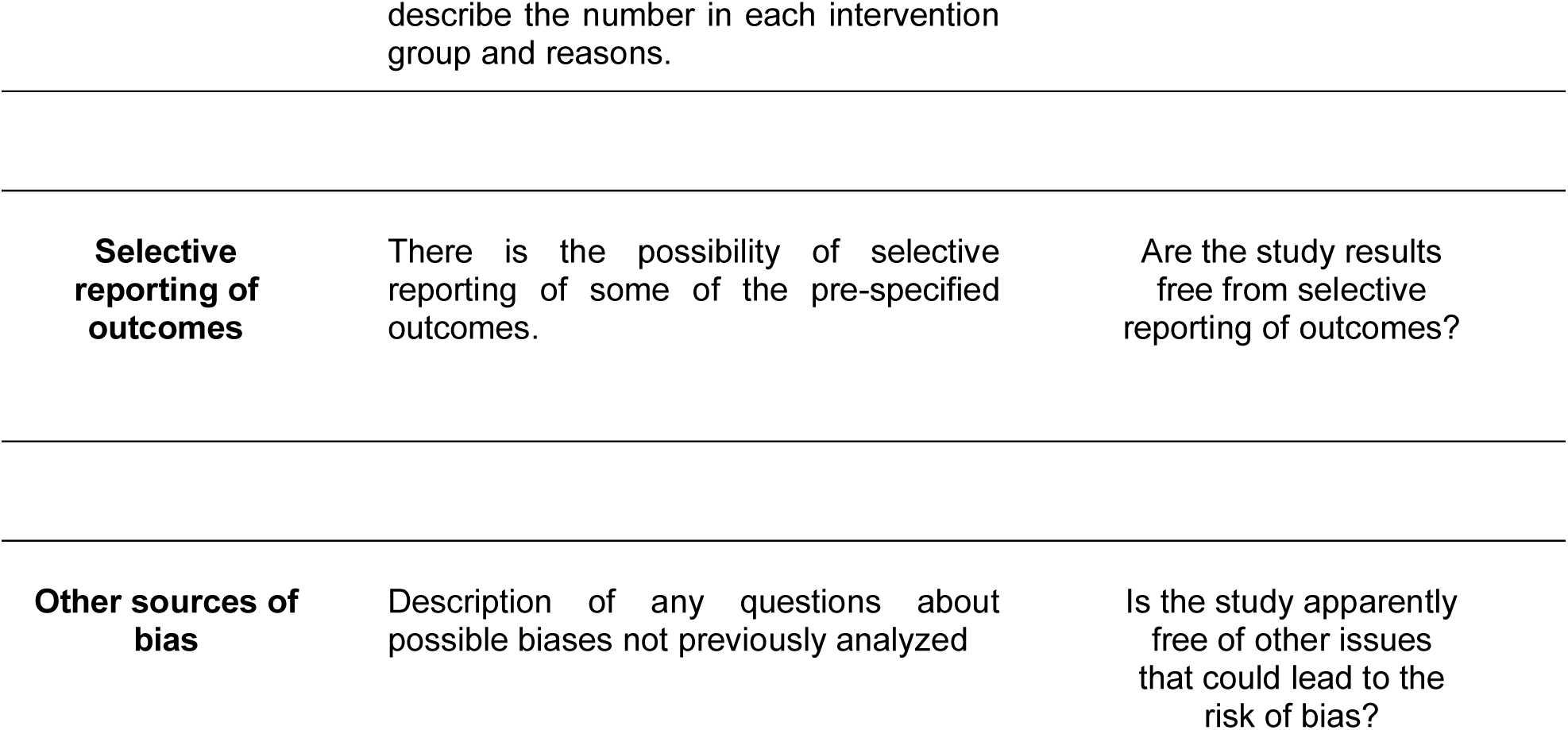
Risk of bias analysis of the Cochrane Collaboration^12^

### Data Collection and Analysis

The citations obtained through the search strategy in the different databases were gathered in a single list, after excluding duplicate citations. The titles and abstracts of all studies were reviewed and those considered potentially relevant were selected for full reading. Those who met the selection criteria were included in the review. The entire study selection process was carried out in pairs, by two independent reviewers.

Both independently extracted relevant data from each study selected for inclusion and compared their findings. For each study, information was collected on the characteristics of the study, participants, interventions and outcomes. The methodological quality of the included studies was also assessed by two independent researchers, in accordance with the recommendations of the Cochrane Handbok (12)

Each RCT received a final score for each of the six domains, according to the overall risk of bias (Table 1), being considered: YES (low risk of bias), UNCERTAIN (uncertain risk of bias) or NO (high risk of bias bias), being:

- Low risk of systematic error or bias: all criteria well described and properly applied;
- Uncertain risk of systematic error or bias: one or more of the first three criteria could not be assessed due to lack of information for the judgment.
- High risk of systematic error or bias: one or more of the first three criteria inappropriately applied.

### Statistical analysis

Data analysis was planned, when possible, for the outcomes of interest between the treated and placebo groups; using Review Manager 5.4 software. (13)

## RESULTS

In June 2021, the search strategy recovered a total of 49 citations, 2 in Cochrane, 2 in PUBMED, 25 in EMBASE, 12 in LILACS and 8 in SCOPUS. After eliminating duplicate citations (n=4), 45 unique studies remained. After reading the titles and abstracts of these studies, 38 were excluded for not meeting the selection criteria and 7 were selected for full reading, after which two met the criteria and were included in this systematic review (Figure 1).

**FIGURE 1:**
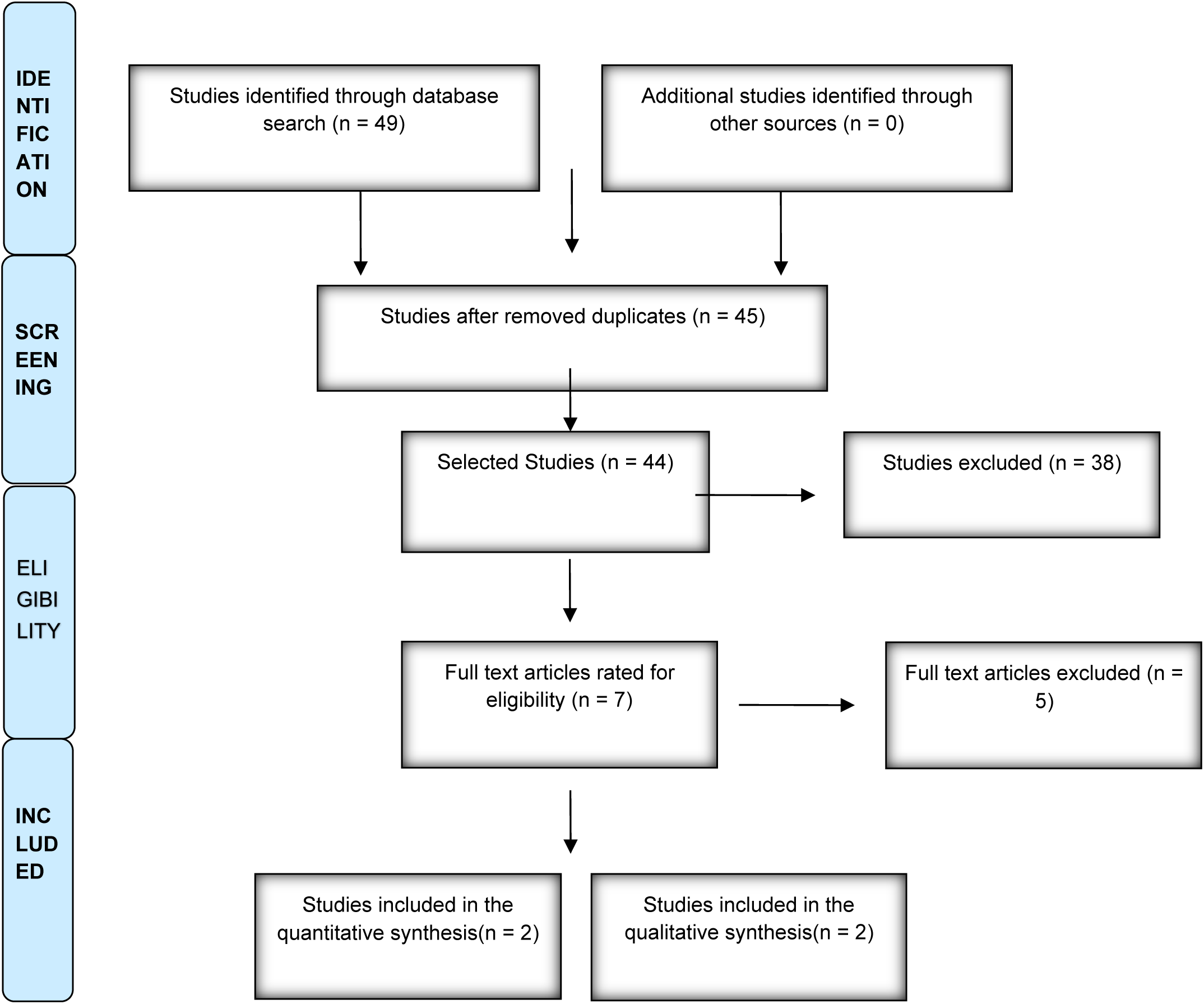
Flowchart of the identification process of studies in electronic databases.

### Included Studies

Two studies (14,15) with a total of 215 participants were included in the systematic review. The main features of these studies are shown in Table 4. The studies were published in 2008 and 2020, respectively, in Turkey and Iran.

### Characteristics of Included Studies

The study by Cingi et al (2008)14 was a double-blind randomized clinical trial (ECRDC) carried out in Turkey and involved 150 patients with a clinical history of allergic rhinitis between June 2006 and February 2007. Patients who were excluded from the study were excluded. had hypertension, diabetes, asthma, drug-induced allergic rhinitis, and acute or chronic rhinosinusitis.

The first group received treatment with spirulina (2000mg/day divided into 5 tablets) for 6 months, and the second group was treated with placebo for the same period.

Before starting treatment and at the end of treatment, patients were evaluated for physical examination findings (shell color, edema, and rhinorrhea), a symptom score such as rhinorrhea, runny nose, nasal congestion and itching, and a diary pertinent to nasal symptoms. (nasal obstruction, runny nose, itching and itching) evaluated 21 times a week. All patients were also submitted to a visual analogue scale, assigning a score from zero to ten to assess the efficacy of spirulina.

Statistical analysis was performed using Student’s t test for independent samples or the Mann-Whitney test. To assess the findings of the physical examination and symptoms found throughout the treatment period, the Wilcoxon test was performed.

There was a loss of 14% of patients, the study was completed with 129 patients divided into 2 groups: intervention consisting of 85 patients and 44 patients on placebo.

The authors described a significant reduction in the score attributed to the numerical scale of symptoms and physical examination findings in the group treated with spirulina for rhinorrhea, runny nose, nasal congestion and itching over time of medication use (p<0.001). When compared to the data collected weekly, the spirulina group had significant efficacy compared to placebo (p<0.001).

The authors did not assess any improvement in the patients’ quality of life. The study by Nourollahian et al (2020)15 was a single-blind randomized clinical trial (ECRMC) carried out in Iran and involved 65 patients with persistent allergic rhinitis from October 2015 to March 2016. Allergic patients were selected from of allergic symptoms (AR signs) and that showed a positive prick test to at least one antigen. Patients with systemic diseases, use of anticoagulants and drugs with anti-inflammatory effects, spirulina intolerance, use of immunosuppressive drugs, pregnant or breastfeeding women and asthmatics were excluded from the study. Patients were divided into two groups: 26 patients treated with spirulina (2000mg/day) for 2 months, and the second group of 27 patients was treated with cetirizine (10mg/day) for the same period. The two groups were compared to each other, before and after treatment. From the sample, 11 patients were excluded for presenting a negative prick test and 1 patient from the group treated with spirulina dropped out of the study.

The patients were evaluated by an analog scale, attributing a score from zero to five, to assess the improvement of symptoms (runny nose, nasal congestion, rhinorrhea, changes in smell and nasal itching) and in relation to factors of the patient’s quality of life (conditions sleep, work and social activities). A blood collection (5ml) was also performed before and after the intervention to assess the levels of interleukins (IL-10, IL-4, IL1, and interferon-gamma).

Statistical analysis was performed using the independent t test and a paired t test. For intergroup comparison, the paired t test was used. A p value < 0.05 was considered statistically significant for all tests.

After therapeutic intervention, symptom improvement was significantly greater in the experimental group (spirulina) than in the control group (citirizine) for all symptoms except nasal itchiness, sneezing, as well as INF-γ (interferon-gamma) levels.

The prevalence of rhinorrhea (P = 0.021), nasal congestion or obstruction (P = 0.039) and decreased smell (P = 0.030) were significantly lower in the experimental group than in the control group; however, no significant change was observed regarding the prevalence of sneezing (P = 0.096) and nasal itching (P = 0.099) between the two groups. Sleep conditions, professional life and social activity improved in both groups, and the improvement was significantly greater in the experimental group. Furthermore, levels of IL-1α (P < 0.001), IL-1β (P < 0.001) and IL-4 (P = 0.008) were significantly lower in the spirulina group compared to those in the control group.

The study showed that a reduction in the severity of all symptoms was observed in both groups after the intervention and these differences were significant for all factors in the spirulina group and for all factors in the cetirizine group except IL-1β (P = 0.109), IL-4 (P = 0.078) and L-10 (P = 0.68).

Table 2 presents the characteristics of the included studies.

**Table 2.**
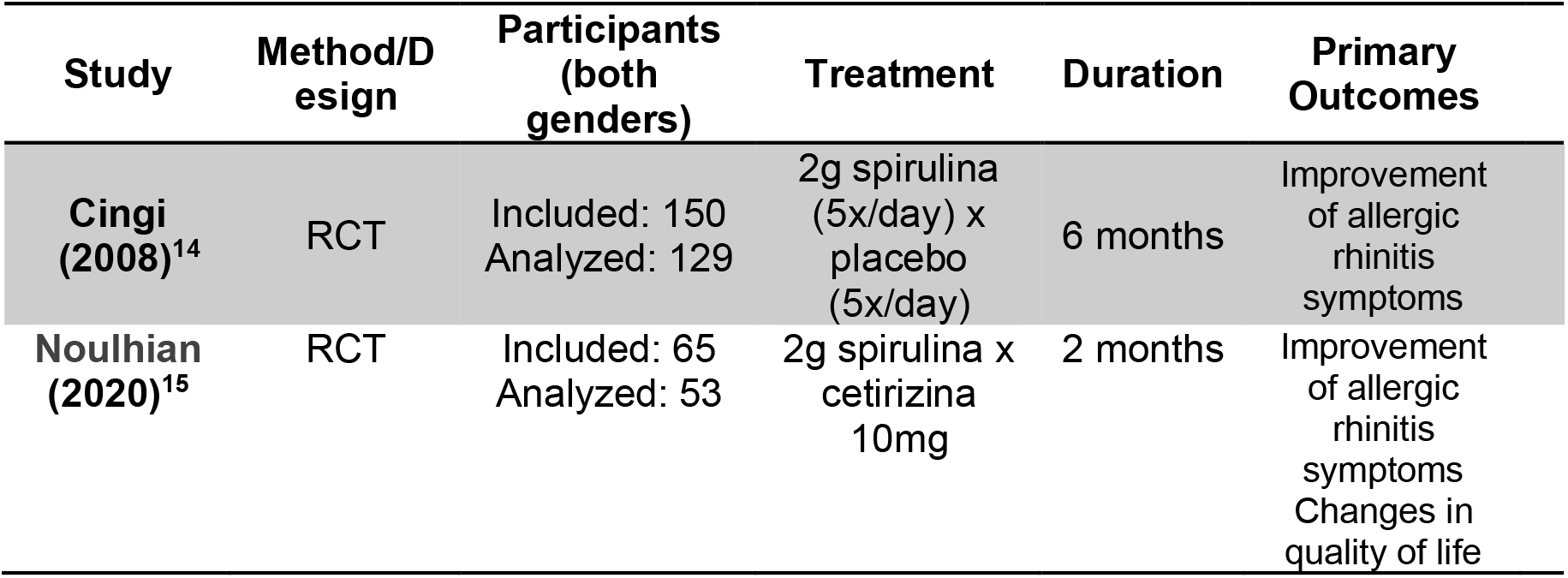
Characteristics of the included Studies.

### Risco de Viés

A Tabela 3 apresenta a análise do risco de viés dos três ECR incluídos nesta revisão sistemática.

**Table 3.**
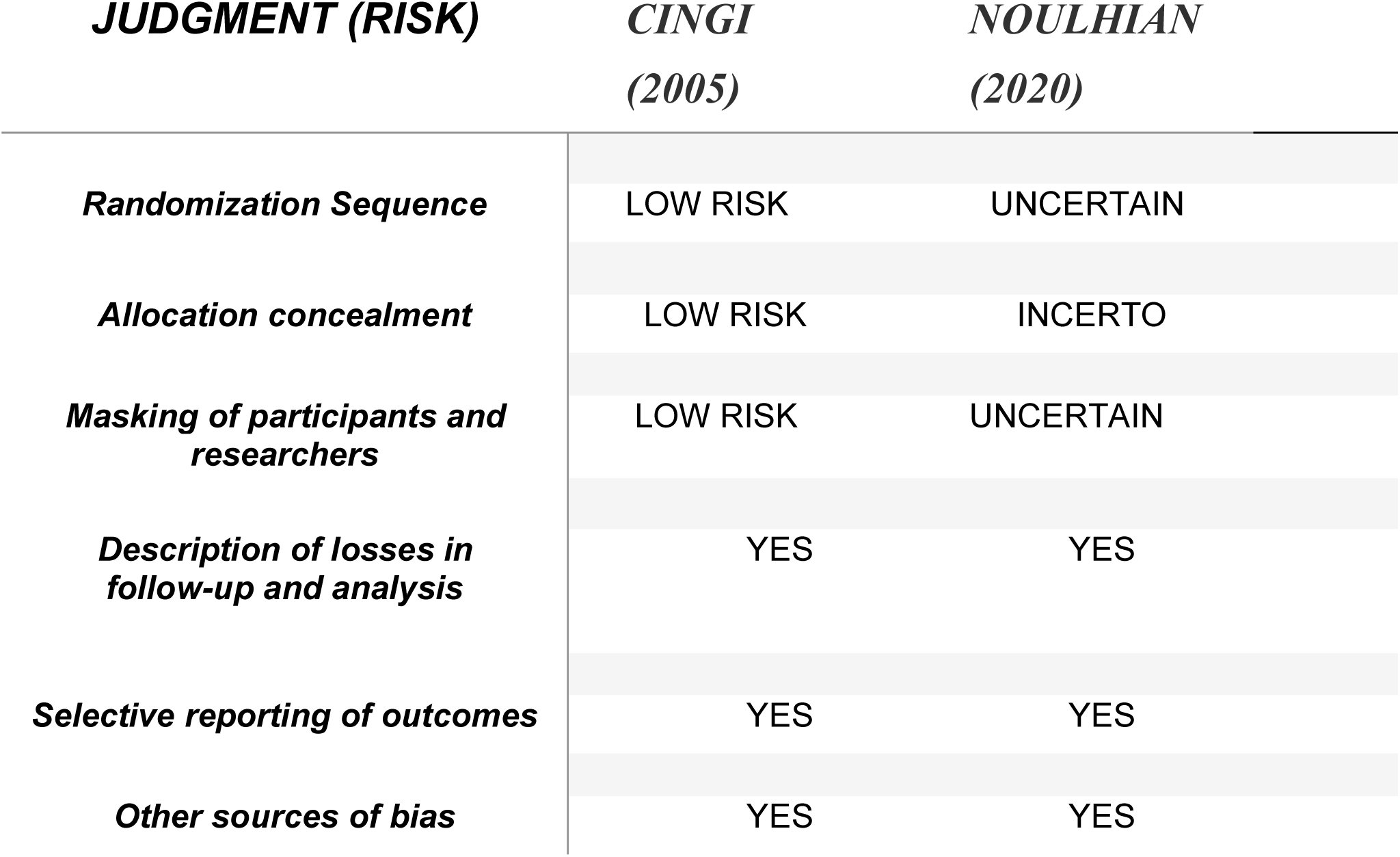
Risk of bias analysis of the included RCTs.

## DISCUSSION

In recent years, alternative therapies have gained popularity for economic reasons and the development of industrialized countries (16). The use of herbs to treat a variety of ailments over the last decade has increased substantially, and often at the recommendation of physicians and often following requests from patients; consequently, the number of RCTs with herbal medicines has increased.

The prevalence of the use of herbs as therapy differs by country and medical condition. The general literature indicates that 25-50% of the population and up to 70% of children have used an alternative medicine (MMA) method at least once (17). A study in Americans demonstrated a 29% prevalence of use of herbal therapy for rhinosinusitis (18). In a study from Spain, 34.4% of 400 patients with allergic diseases (allergic rhinitis, asthma and atopic dermatitis) used at least one type of alternative medicine method and, among these patients, 31.5% had already used it. natural medicine (19). A study in England showed that 65% of respondents had used MMA for rhinosinusitis or nasal polyposis (20). A study from Israel reported a 19% rate for the use of MMA to treat rhinosinusitis (21).

Given the widespread and growing use of herbal therapy and its potential pharmacodynamic and pharmacokinetic effects, an understanding of its use for various conditions may be beneficial.

Allergic rhinitis is a chronic inflammatory condition caused by an exaggerated immune system response to common allergens. Most drug therapies tend to be palliative and, in some cases, are associated with adverse effects.

Mao et al. (2005) evaluated the therapeutic application of the spirulina supplemented diet for allergic rhinitis by measuring the production of cytokines important in the regulation of the IgE-mediated allergy response (24). The study reported that spirulina significantly reduced IL-4 secretion by 32% only when patients received 2000 mg/day.

This systematic review of the literature identified only two RCTs, which assessed the efficacy of spirulina for allergic rhinitis, taking a group of patients treated with placebo or cetirizine as a control.

A strength of this review was its methodological rigor, following the Cochrane Collaboration recommendations, including a sensitized and unrestricted language search, the involvement of two independent investigators in study selection, data extraction, and quality assessment of included studies. One limitation was the fact that it did not search for unpublished studies on the subject, possibly described in abstracts of conferences, symposia and scientific conferences.

As a food supplement, spirulina was easily accepted by patients. The compliance and adherence of patients is quite high (25). More than that, the diaries kept by the patients revealed a statistically significant positive effect when compared to the placebo.

Karkos et al. (25) concluded that there is evidence for the positive effects of spirulina on AR, although further testing is needed in this regard. In addition to human studies, animal research has also examined the effect of spirulina platensis on IL-4 and IFN-γ expression in the serum of a laboratory rat affected with RA (26,27). They concluded that spirulina platensis is very effective in treating RA by regulating the expression of IL-4 and IFN-γ, as well as adjusting the imbalance in the Th1/Th2 cytokine network.

These findings were also confirmed by a recent study that showed spirulina had anti-inflammatory properties and inhibited histamine release from mast cells. (26.27). The present study on the effect of spirulina in patients with RA confirmed this effect in humans. Given the results of several studies carried out in rodents, as well as the long and historical use of spirulina by humans, the Food and Drug Administration has approved its safety (28). Therefore, there are no concerns regarding the administration of spirulina.

On the other hand, in addition to its therapeutic application, spirulina can have effects as a nutritional and pharmaceutical supplement. In previous studies, spirulina was shown to contain high levels of proteins, essential fatty acids, essential amino acids, micro and macrominerals, vitamins and polysaccharides; thus offering a favorable combination of all the components needed by the human body (29-31).

It should be noted that, considering the nature of AR and the effect of exposure to allergens on the severity of clinical symptoms, different environmental conditions can affect the results. However, by conducting a randomized study and selecting an appropriate time frame, we tried to reduce the impact of seasonal factors on the severity of allergic symptoms.

Spirulina treatment has a major disadvantage in terms of the high dose required; a patient needs to take 2 g of spirulina equivalent to 4 to 5 capsules daily. In practice, many patients can resist taking such a high volume of medication.

Ideally, recommendations regarding the use of these interventions should involve a shared decision-making process between the healthcare provider and the patient, and the potential effectiveness, risks and benefits, and financial implications of their use should be taken into account. More large-scale studies are needed to fully understand the role health supplements can play in controlling this condition.

The treatment of allergic rhinitis involves the proper diagnosis and the standardization of questionnaires can effectively contribute to the adequacy of each situation to the therapeutic reality. It is worth noting that in recent decades, the therapeutic approach to allergic rhinitis has undergone changes in a similar proportion to new discoveries in the area.

The subject of this systematic review, spirulina was evaluated in only two placebo-controlled studies. The number of participants in these studies was small. The differences between the included studies made statistical processing in meta-analysis impossible. Heterogeneity has been a problem for carrying out meta-analysis and it is suggested that new studies be carried out following parameterization to describe the results, as outlined in CONSORT (Consolidated Standards Of Reporting Trials). This shows limitations in the interpretation of the effectiveness of the treatment of allergic rhinitis with spirulina.

In this context, given the very low level of evidence, it is essential to carry out new RCTs involving spirulina therapy for patients with allergic rhinitis, and other parameters should be considered by researchers in light of recent scientific discoveries. Therefore, further studies with a longer follow-up period are recommended to investigate the drug’s effect on the relapse of clinical symptoms and to assure patients that there are no side effects of spirulina.

## CONCLUSION

The studies included were in favor of the use of spirirulina, but the level of evidence is very low and limited and it is suggested that new RCTs be carried out, it is suggested that new studies be carried out following parameterization for description of results, as outlined in CONSORT.

## Data Availability

All data produced in the present study are available upon reasonable request to the authors

https://pubmed.ncbi.nlm.nih.gov/18343939/

https://pubmed.ncbi.nlm.nih.gov/32773785/

